# SARS-CoV-2 live virus culture and sample freeze-thaw stability

**DOI:** 10.1101/2023.07.29.23293373

**Authors:** Phyllis J. Kanki, Donald J. Hamel, Stefan Riedel, Sanjucta Dutta, Annie Cheng, Charlotte A. Chang, Ramy Arnaout, James E. Kirby

## Abstract

The COVID-19 pandemic has presented unique diagnostic challenges including the need to store and test large number of samples for clinical and research studies. While SARS CoV-2 diagnosis relies on RT-qPCR and antigen testing, live virus culture remains an important surrogate for viral “infectiousness”, as we previously described in “SARS-CoV-2 Antigen Tests Predict Infectivity Based on Viral Culture: Comparison of Antigen, PCR Viral Load and Viral Culture Testing on a Large Sample Cohort” (Clin Microbiol Infect, 2022, <u>PMC9293398</u>). Live virus isolation and characterization has also been important to the SARS CoV-2 research community, to assess viral fitness, cellular tropism, and live virus neutralization, particularly with the emergence of new variants. Many clinical and research studies make use of samples that are frozen in transport media and investigated at later dates. The effect of freezing on RT-qPCR results is well established. However, the effect of freeze-thaw on viral viability has not been. Here, we therefore examined the effect of freeze-thaw on viral culture isolation from a large number of clinical samples that were split, and then cultured either fresh or after being frozen for 7 or 17-18 days. Samples represented the range of viral loads (genome copies/mL) observed in our patient population. We found that freeze-thaw did not significantly affect viral culture isolation. Therefore, the ability to assess infectiousness of samples previously frozen in transport medium appears to be maintained.

---

We previously reported on severe acute respiratory syndrome coronavirus 2 (SARS-CoV-2) antigen tests and their ability to predict infectivity based on viral culture (1) including Omicron and Delta variant analyses (2). The detection of infectious virus, also referred to as live virus or replication-competent virus, by virus culture is demonstrated by in vitro infectiousness on cell lines and is regarded as an informative surrogate of viral transmission. While numerous studies have reported on the influence of storage and freeze-thaw conditions on qPCR quantitation of SARS-CoV-2, the impact on detection of infectious virus has been infrequently reported. In one study, 5 viral isolates from 16 specimens underwent a single freeze-thaw cycle; the study reported no significant difference in rate of virus isolation, but samples that underwent freeze-thaw took a longer time to detect by culture (13.8, SEM=1.91 versus 4.28, SEM=0.39 days, respectively; P=0.0001) (3).

We therefore performed a larger study to confirm that freezing of nasopharyngeal samples in viral transport medium did not affect viral viability. Nasopharyngeal swabs were obtained in 3 mL of saline or viral transport medium at COVID-19 testing sites affiliated with our institution in Boston, MA from March through June 2021 (1). After diagnostic qPCR testing, 31 de-identified PCR-positive samples, selected for analysis solely based on viral load distribution, were aliquoted; one aliquot was cultured immediately and two were frozen at -80°F and then thawed after 7 or 17-18 days, respectively, and similarly cultured. Identically treated, PCR-negative respiratory samples were included as controls (n=4) with each batch of cultured samples. Human subjects research was approved by our respective Institutional Review Boards.

Viral culture in Vero E6 (ATCC CRL-1586) cells was performed as described previously (1). SARS-CoV-2 real-time (RT)-qPCR testing of samples and Vero cell culture supernatants was performed using the Abbott Molecular M2000 Real-Time or Alinity m SARS-CoV-2 assays with limits of detection (LoD) of ∼100 genome copies/mL (4, 5). On days 3, 6, and 13-14 days of culture, equal volumes of cell culture supernatant and VXL buffer (QIAGEN, Germantown, MD) were combined for subsequent nucleic acid extraction and SARS-CoV-2 RT-qPCR. Samples from which at least two of three sequential supernatant viral loads exceeded the LoD were considered culture positive (6).

With results from fresh sample aliquots as the reference standard, sensitivity and specificity of viral culture isolation from 7-day frozen aliquots was 100% (95%CI, 88.8%-100%) and 100% (95%CI, 39.8%-100%), respectively, for the 31 qPCR-positive and 4 qPCR-negative samples examined. For 17-day frozen sample aliquots, sensitivity and specificity were 93.5% (95%CI, 78.6%-99.2%) and 100% (95%CI, 39.8%-100%), respectively. Of the two discordant samples from the 17-day frozen samples, one had negative culture supernatant on days 3, 6, and 14; the other was positive on day 3 but negative on days 6 and 14.

Our validation study demonstrated no impact of respiratory sample freezing on isolation of infectious virus from samples with >5.3 log_10_ viral load (Fig. 1). These results suggest that cryopreserved respiratory samples can be reliably utilized for further study and characterization of infectious SARS-CoV-2.

**Figure 1.**
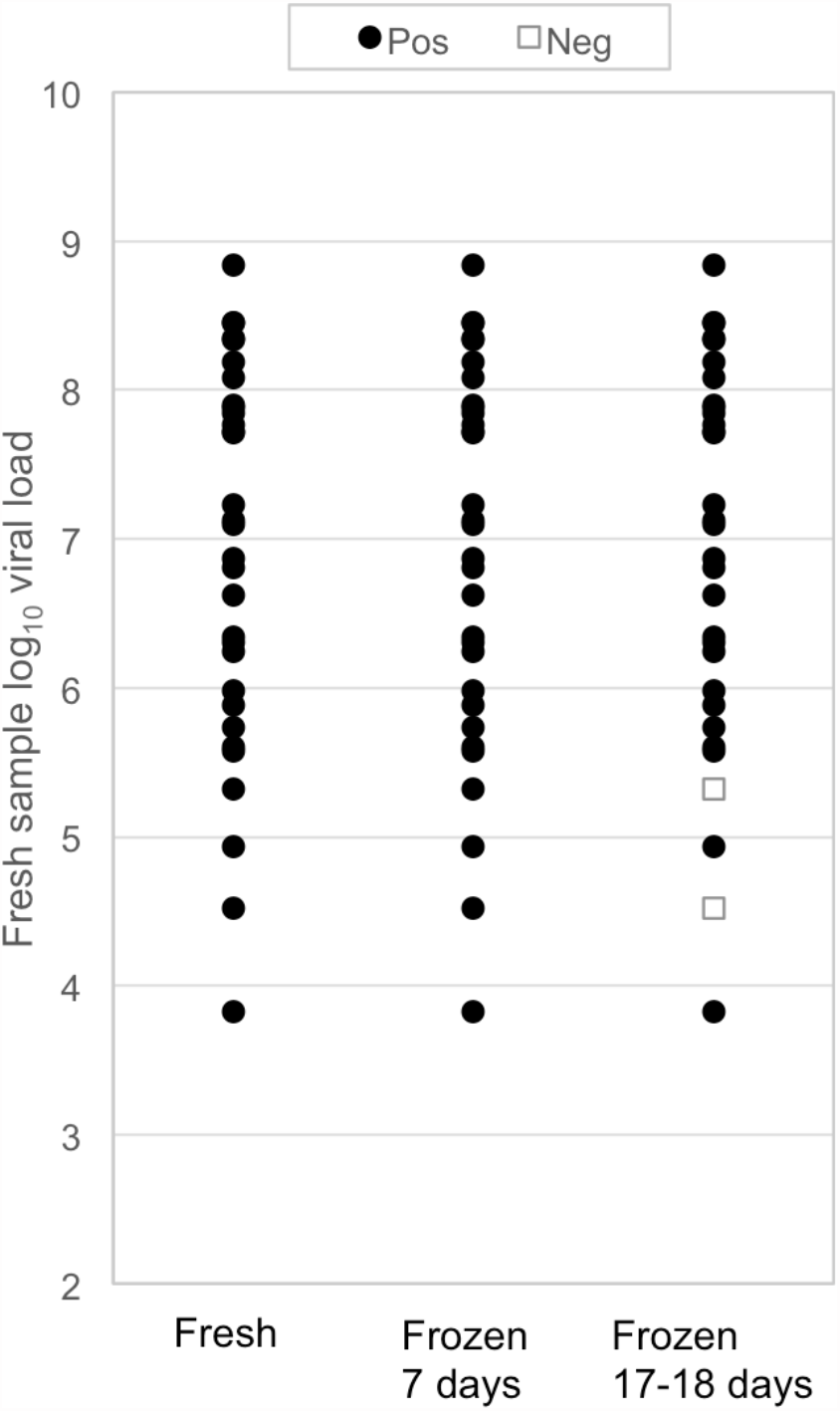
Detection of viable SARS-CoV-2 virus in patient samples with different viral loads split into three aliquots and cultured in Vero cells either immediately or after being frozen for 7 or 17-18 days. Culture viability and sample viral loads were determined as described in methods description and previously (1, 7). Filled circles, detection of culture viable virus; open squares (positive), viable virus not detected (negative). Of the 31 qPCR positives samples, there was no loss of SARS-CoV-2 culture viability after being frozen for 7 days, and a non-significant loss of viability after being frozen for 17-18 days (McNemar test *P*=0.1573).

## Data Availability

All data produced in the present study are available upon reasonable request to the authors

